# The YUVAAN cohort: an innovative multi-generational platform for health systems and population health interventions to minimize intergenerational transmission of non-communicable diseases in India

**DOI:** 10.1101/2023.08.30.23294810

**Authors:** Demi Miriam, Rubina Mandlik, Vivek Patwardhan, Dipali Ladkat, Vaman Khadilkar, Neha Kajale, Chidvilas More, Ketan Gondhalekar, Jasmin Bhawra, Tarun Katapally, Anuradha Khadilkar

**Affiliations:** Hirabai Cowasji Jehangir Medical Research Institute (HCJMRI), Jehangir Hospital, Pune, Maharashtra, India; CHANGE Research Lab, School of Occupational and Public Health, Faculty of Community Services, Toronto Metropolitan University, Toronto, ON, Canada; DEPtH Lab, School of Health Studies, Faculty of Health Sciences, Western University, London, ON, Canada; Interdisciplinary School of Health Sciences, Savitribai Phule Pune University, Pune, Maharashtra, India

**Keywords:** non-communicable diseases, health systems, multi-generational cohort, socioecological determinants of health, intergenerational inequities, life course, climate change, digital health

## Abstract

**Introduction:** Non-communicable diseases (NCDs) pose a significant health burden in India, with preventable risk factors contributing to their prevalence. Intergenerational inequities can exacerbate the transmission of health risks to further disadvantage vulnerable populations. Taking a life course perspective, this multi-generational cohort study aims to investigate behavioural, socio-ecological, and socio-economic determinants of growth and NCD risk, as well as healthcare access and utilization among rural households that include preadolescent children and their parents.

**Methods:** The study is being implemented by Hirabai Cowasji Jehangir Medical Research Institute (HCJMRI) utilizing a prospective multi-generational cohort design to investigate NCD risk across 15 years. Data are being collected from 12 villages around Pune, Maharashtra, India. The primary population enrolled includes apparently asymptomatic (i.e., healthy) children aged 8 to 10 years and their parents.

The sample size calculation (N=1300 children) for this longitudinal prospective cohort was driven by the primary objective of assessing trajectories of growth and NCD incidence across generations. A total of 2099 children aged 6 to 10 years have been screened since April 2022, of whom 1471 have been found to be eligible for inclusion in the study. After obtaining informed consent from parents and their children, comprehensive bi-annual data are being collected from both children and parents, including clinical, behavioural, healthcare access and utilization as well as socio-ecological and socio-economic determinants of health. Participants (children and their parents) are being enrolled through household visits, and by arranging subsequent visits to the primary health facility of HCJMRI. Clinical assessments include anthropometric measurements, blood samples for a wide range of NCD indicators, bone health, and muscle function. The long-term data analysis plan includes longitudinal modeling, time-series analyses, structural equation modeling, multilevel modeling, and sex and gender-based analyses.

Ethics approval has been obtained from the institutional ethics committee, the Ethics Committee Jehangir Clinical Development Centre Pvt Ltd. Written informed consent is obtained from adults and written informed assent from children.

**Discussion:** As of May 2023, 378 families from 10 villages have been enrolled, including 432 preadolescents and 756 parents. Preliminary results not only highlight the double burden of malnutrition in the cohort with undernutrition and overweight/obesity coexisting among children and parents, respectively but also identify high rates of diabetes and hypertension among adults in rural areas. Findings can inform the development of targeted interventions to reduce NCDs, address intergenerational health inequities, and improve health outcomes in vulnerable populations.

## INTRODUCTION

Non-communicable diseases (NCDs) pose an immense burden on global health, with approximately one in three adults living with more than one chronic condition (1). According to the World Health Organization, NCDs are responsible for approximately 71% of all global deaths, which is equivalent to around 41 million people per year (2). Among the complex myriad of factors that influence NCD risk (3), maternal health and social circumstance (i.e., growing up in poverty) can initiate a cycle of intergenerational transmission of health risks which further disadvantage marginalized and vulnerable populations (4–6).

Intergenerational inequities have particularly negative implications for children’s health in low and middle-income countries where a significant proportion of the population can be classified as low socioeconomic status (7,8). Mortality and morbidity rates are significantly higher among children born in low-income families (9,10). Moreover, social determinants of health, such as poverty and low education, can pass from one generation to the next and continue to adversely impact the health status of disadvantaged populations.

In understanding, and potentially preventing intergenerational transmission of health risks, it is critical to take a life course perspective (11), with a particular emphasis on the vital phases during which the growth and development of children are determined (12–14). Health systems and population health interventions that minimize risk transmission require a multi-generational approach (14) that is intersectoral, integrated, and comprehensive (15). A multi-generational approach to reducing health inequities is imperative for transformative change in the global south to reverse the historical injustices of colonialism (16) that continue to drive intergenerational transmission of health risks due to pervasive economic, political, and social inequities (17,18). In an effort to minimize intergenerational transmission of health risks, a focus on countries such as India is essential due to its significantly large share in global population (19,20). However, given the broader context where issues such as climate change and communicable disease pandemics challenge global health systems, a multi-generational approach is not enough, as the wider socio-ecological determinants of health (21) identified in the United Nations Sustainable Development Goals must be addressed taking a holistic health perspective (22).

Thus, in taking a multi-generational approach to address global health inequities and enhance health outcomes, it is crucial to understand how socioecological determinants of health influence children’s health in countries like India, where children face significant challenges in early years due to malnutrition and food insecurity (23–27) – a health inequity, which can widen due to climate change impacts on food security (28,29). More importantly, intergenerational transmission of NCD risks needs to be mitigated through the development and implementation of health systems and population health interventions during critical phases of childhood (30,31).

Yet, from a multi-generational perspective, parental health and socioeconomic status also have direct impacts on children’s health (32), particularly in terms of nutrition and growth (33). This parental influence, combined with the critical phases of preadolescence and adolescence in shaping both pubertal growth spurt as well as NCD-related health behaviours such as appropriate nutrition and physical activity (30,34), reiterate the need for a multi-generational approach in maximizing growth and minimizing NCD risks in adulthood.

Still, there is little longitudinal evidence in mitigation of intergenerational transmission of NCD risk, particularly among disadvantaged communities in rural areas in low– and middle-income countries such as India (35). To address these gaps, the YUVAAN (YoUng adolescents’ behaViour, musculoskeletAl heAlth, growth & Nutrition) multi-generational platform was conceptualized to understand and influence trajectories of growth and NCD risks among rural preadolescents (8-10-year-old children) and their parents over a period of 15 years. YUVAAN’s overall goal is to not only capture the broad range of socioeconomic and socioecological determinants of health across generations, but also to understand healthcare access and utilization among a rural cohort – evidence that currently does not exist in India.

To achieve this overall goal, YUVAAN has the following objectives: **1)** to understand trajectories of growth among rural preadolescents (aged 8-10 years) and their parents over a period of 15 years, **2)** to investigate intergenerational patterns of NCD health risks and growth (including muscle and bone health) among preadolescents (8-10-year-olds) including the health of their parents and future offspring in rural areas, **3)** to identify and address socioeconomic and socioecological determinants of health (i.e., climate change impacts, sociocultural factors) associated with growth trajectories and intergenerational NCD risk, and, **4)** to strengthen community-based, social and behaviour change communication strategies and implement interventions that improve the nutritional and health status of preadolescents and their parents by exploring and improving their knowledge, attitude, and practices.

## MATERIALS AND METHODS

### Ethics Approval & Informed Consent

Ethics approval for this study was obtained from the Institutional Ethics Committee (Ethics Committee Jehangir Clinical Development Centre Pvt Ltd). The study has also been registered on the clinicaltrials.gov portal (ClinicalTrials.gov Identifier: NCT05603793). All data are collected after obtaining written informed consent and informed assent from adults, and written informed assent from the children.

### Study Design and Setting

YUVAAN is a longitudinal, prospective, multi-generational cohort study set in the rural areas (as defined by the Census of India) of the Pune district within the Western Indian state of Maharashtra (36). A total of 22 villages were screened between January and June 2022, with 12 meeting the inclusion criteria described in (**Figure 1**). According to the Census of India, a place is considered to be a village if it meets the following criteria: **1)** it has a population of less than 5,000 individuals; **2)** the majority of the male workforce is engaged in agricultural activities, and **3)** the population density is less than 400 individuals per square kilometre (36,37).

**Figure 1.**
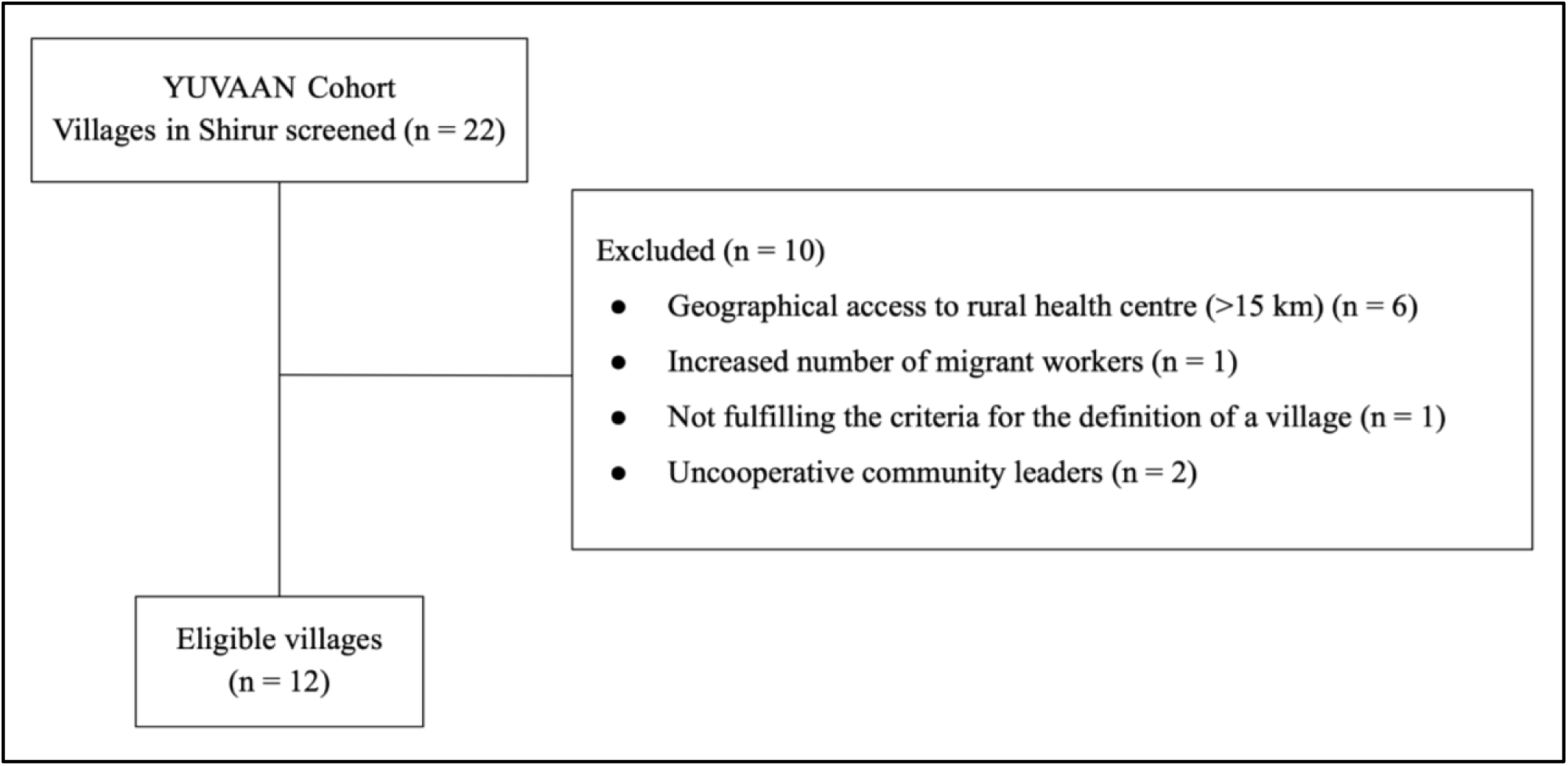
– Study eligibility criteria.

As illustrated in **Figure 2**, the villages chosen are located on either side of the National Highway, NH 753F, which passes through Ranjangaon, the location of the Hirabai Cowasji Jehangir Medical Research Institute’s (HCJMRI) rural centre – the central hub for implementation and evaluation of YUVAAN. The centre has clinical expertise in growth, nutrition, paediatric endocrinology, and musculoskeletal health, a multidisciplinary team including paediatricians, paediatric endocrinologists, nutritionists, community social workers and psychologists, and houses relevant infrastructure (i.e. Dual X-ray Absorptiometry [(DXA]), pQCT [(Peripheral Quantitative Computed Tomography], etc) to measure height, assess body composition, bone mineral content, bone mineral density, muscle mass and function, grip strength, and bone age of patients (38).

**Figure 2.**
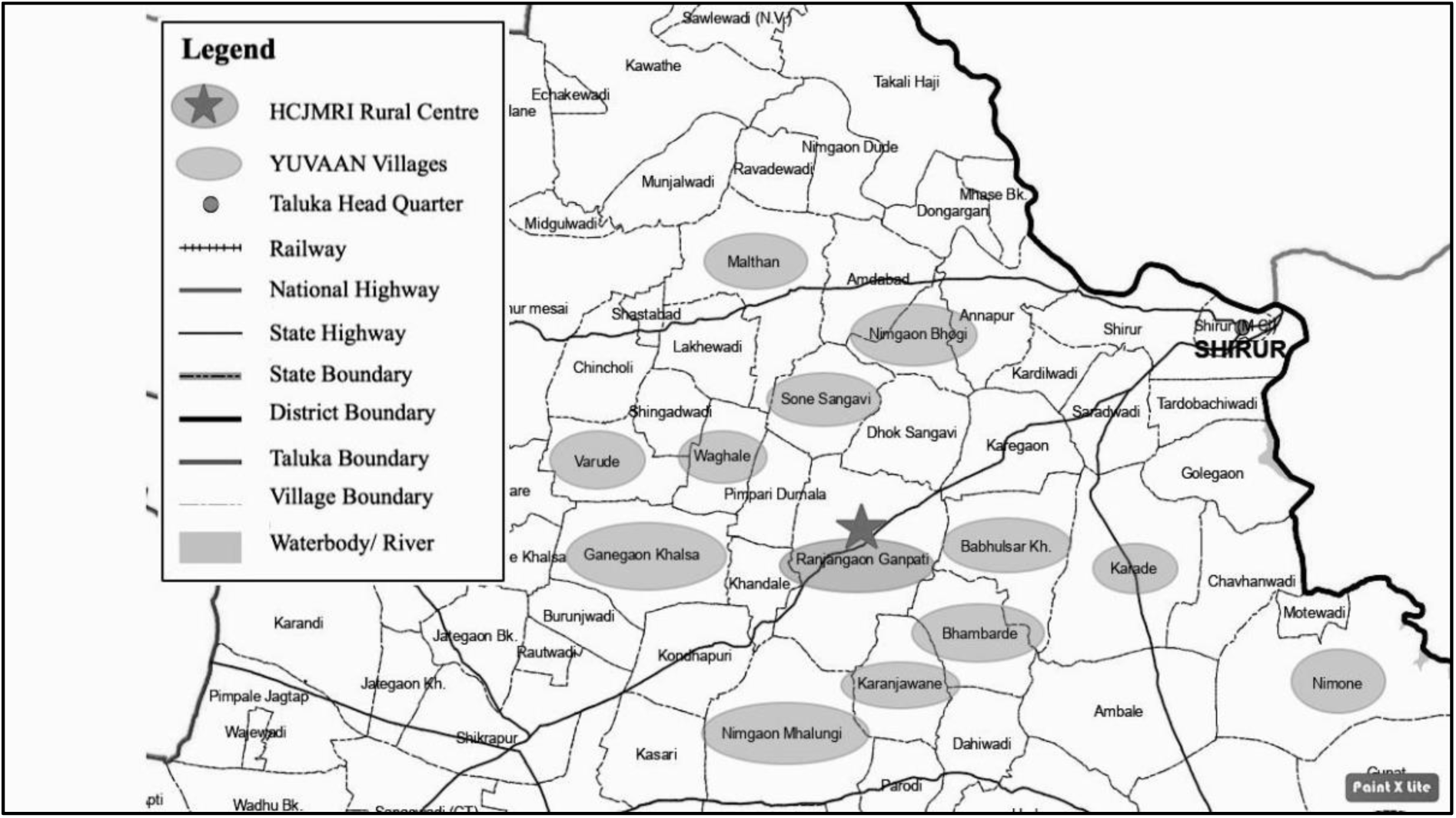
– Map of the study site and included villages.

The 12 selected villages have an average population of 2271±646 people. Before data collection from households in the villages, our team of field workers visited each village to interview the administrative head (Sarpanch) and obtain demographic details about the village which the Sarpanch collected from official data registers. The interview collected information on village population, primary employment sector, number of families residing in the village, percent of migrant population, preferred healthcare choice for medical problems, accessibility of the primary health centre, and designated points of contact for health-related matters.

### Study Participants

The primary study units are households comprising preadolescent boys and girls aged 8 to 10 years, as well as their parents **(Figure 3).** <u>Inclusion criteria:</u> participants are considered eligible if they: are apparently healthy (i.e., asymptomatic) preadolescents aged between 8 and 10 years during the period of enrolment; have both apparently healthy (i.e., asymptomatic) biological parents (mother and father) present during the period of enrolment; have parents in the average range of adult height with reference to national representative sample (Indian men: 177±2SD cm; Indian women: 162±2SD cm, National Family Health Survey-V). <u>Exclusion criteria</u>: participants are excluded if they: have any chronic untreated/progressive condition that would adversely affect growth, bone and/or muscle health of the children; are migrant families or temporary residents as defined by the Census of India (39); or if parents or children refuse to provide informed consent. To ensure a fair opportunity for eligible families to participate in the cohort study, a maximum of 2 children are being enrolled in each family. The general procedure for recruitment is as follows: after obtaining permission from local education and health administration, a list of prospective preadolescents is collected from each village head’s office. School health visits are then conducted to screen children based on the inclusion and exclusion criteria. Parents of prospective children are then approached to determine their interest in the study. **Figure 3** shows the comprehensive plan of current and future enrollment across generations.

**Figure 3.**
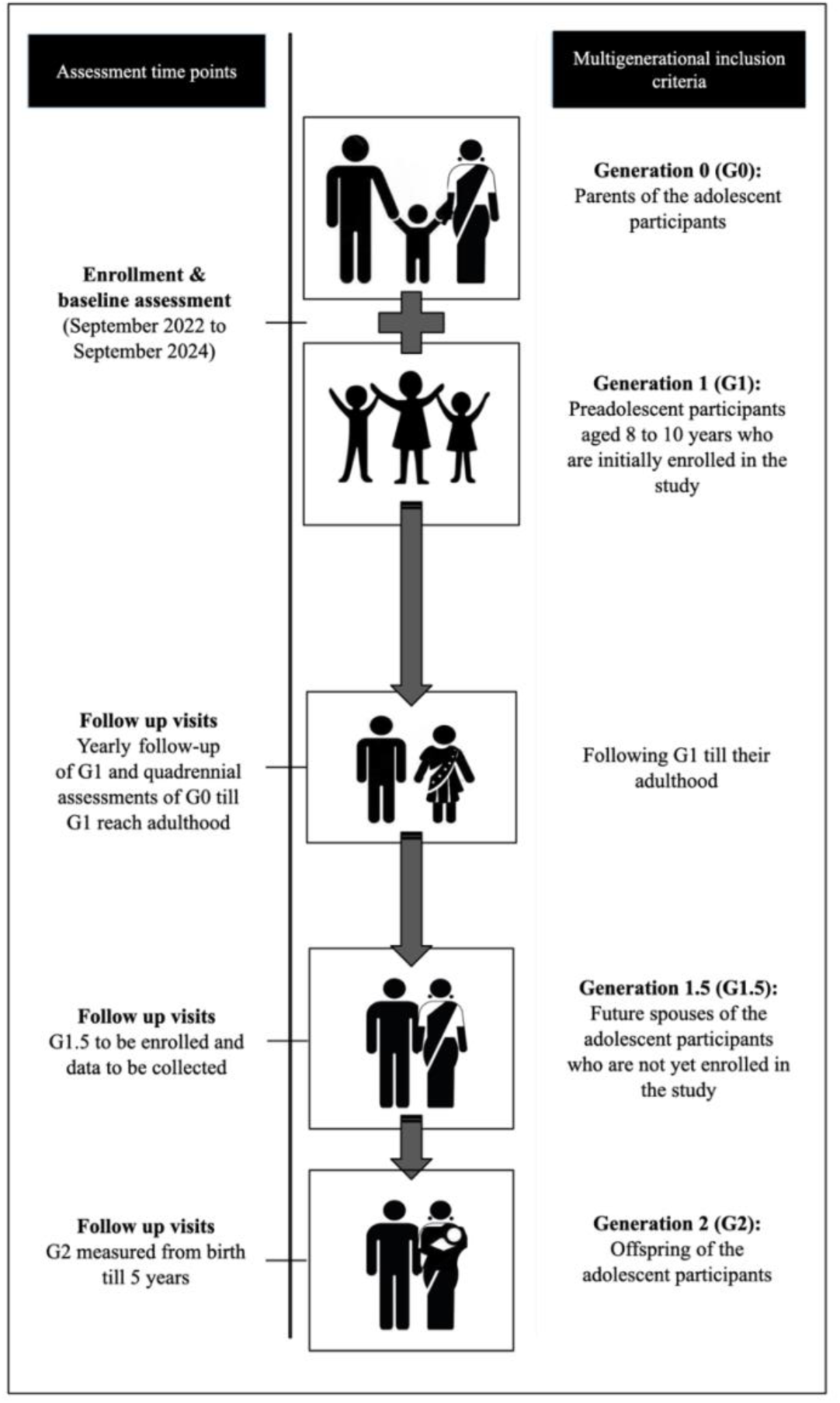
– The flow of assessments targeting the multi-generational cohort.

### Sample Size

While this multi-generational longitudinal study aims to capture a comprehensive range of social determinants, individual behaviours, clinical indicators, and NCD risk outcomes, to determine sample size, growth trajectory of preadolescents was prioritized to capture changes over time. Since height is recognised as a good indicator of a child’s growth and depicts cumulative linear growth, reference data on height-for-age was used from the Indian population (40) to determine the sample size. The software package G*Power ver. 3.1.9.7 was used to calculate the sample size (41). Cohen’s D effect size was found to be d = 0.01 with a standard deviation (SD) of 4 cms in the heights of males (mean=153 cms) and females (mean=148 cms). With a 20% expected dropout rate, a total sample size of 1300 was calculated at a 95% significance level and 80% power with a small effect size of f = 0.02.

### Data Collection

As shown in **Figure 4**, longitudinal data for the YUVAAN study is being collected on physical health, behavioural health, healthcare access, and socioecological and socio-economic factors. A detailed timeline of data collection and assessments is presented in **Supplementary Material (Tables 6, 7, and 8).**

**Figure 4.**
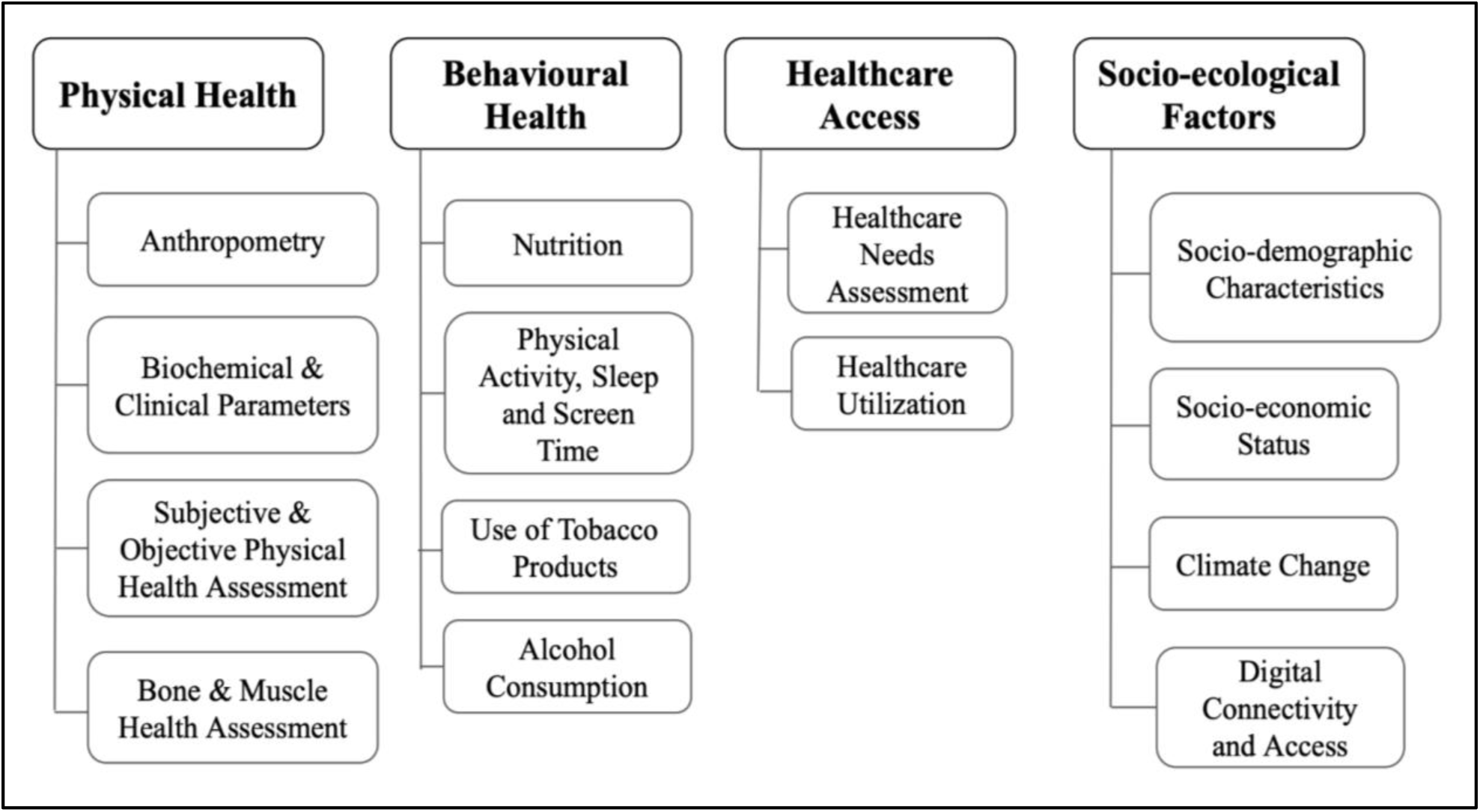
– Data collection variables.

### Physical Health

The following measures will be collected from preadolescents and parents.

***Anthropometry (objectives 1 and 2)***: This includes measurement of height, weight, sitting height, waist circumference (for parents & children) and mid-upper arm circumference (MUAC) (for children). Instruments used for measurement include a portable stadiometer to measure height and sitting height (Seca 213 Portable Stadiometer; Seca, Hamburg, Germany), Tanita Body Composition Analyzer (Model MC-780 MA; Tanita Corporation of America, Arlington Heights, Illinois) to measure weight, and stretch-resistant Seca tape (Seca 201) to measure waist circumference and MUAC. Height for age (HAZ), weight for age (WAZ), and body mass index (BMI) for age (BAZ) Z-scores for children are being computed using ethnic-specific growth references (42). Categorization of primary participants into normal weight, thin, or obese is also being done based on MUAC using Indian reference data (43).

***Biochemical & Clinical Parameters (objectives 1 and 2)***: **Serum vitamin D** [by enzyme-linked immunosorbent assay (ELISA) using standard kits (DLD Diagnostika GmbH (Hamburg, Germany); intra-assay coefficient of variation (CV): 5%; interassay CV: 7.8%)], **parathyroid hormone (PTH)** [by the ELISA technique using standard kits (Biomerica Inc (Irvine, California); sensitivity: 0.17 pmol/L; interassay CV: <4%; intra-assay CV: 3%-6%)], and **luteinizing hormone (LH)** [by the Chemiluminescent Microparticle Immunoassay (CMIA) method using the ARCHITECT LH Reagent Kit (6C25)] for children; **fasting blood sugar** [glucometer], **haemoglobin** (for parents & children) [on the Horiba Yumizen H500 by spectrophotometry], and **lipid profile** for parents and **indicators of bone metabolism and growth modulators such as serum calcium, phosphate, alkaline phosphatase** for children [on the Selectra Pro S Automatic Biochemistry Analyzer using the QLine BioTech (India) Rapid Test Kit].

***Subjective and Objective Physical Health Assessment:*** Medical History comprising fracture history, history of surgeries and medical conditions and history of consumption of or allergies to any medications or supplements are gathered using clinician-administered surveys to address study **objectives 1, 2, 3 and 4**. Reproductive History (for females) comprising menstrual history for the primary participant and her mother and obstetric history for the mother are collected using a questionnaire **(objectives 2 and 4).**

***Bone and Muscle Health Assessment:*** Physical fitness parameters reflecting muscular strength, power, and force are assessed for parents & children using the Leonardo Mechanograph Ground Reaction Force Plate (Novotec Medical, Pforzheim, Germany) **(objectives 1 and 2)**. Body composition, whole-body and volumetric bone mineral density (BMD) and bone mineral content (BMC) are measured for parents & children using the GE Lunar iDXA (Wisconsin, MD) (Software encore version 16) and the Stratec XCT 2000 pQCT equipment (Stratec Inc., Pforzheim, Germany) **(objectives 1 and 2).**

### Behavioural Health

The following measures will be collected from preadolescents and parents.

***Nutrition***: Dietary data are being collected by the 24-hour dietary recall, multiple pass method over 2 days – one weekday and one weekend day (Saturday or Sunday). This enables us to observe trends in dietary patterns, food security, and the influence of climate change (parents & children) (44) **(objective 3).**

***Physical Activity, Sleep, and Screen Time***: Information collected about physical activity comprises activities listed in the questionnaire that are participant-specific i.e., occupational activities have been listed for the adults and more play-related activities have been listed for children. There are options to note additional activities if the participant engages in them and these are classified as light, moderate, or vigorous by a trained Research Associate based on the MET (Metabolic Equivalent of Task) scores. Data related to the time spent performing the activity, including sleep and time spent watching screens (television / mobile screen) during a weekday and a weekend day (Saturday or Sunday) are also collected **(objectives 1 and 2)**.

***Use of Tobacco Products:*** Information is being gathered on the use of tobacco products among the participants. This includes collecting data on smoking habits, such as the frequency and quantity of cigarettes or other tobacco products consumed. Additionally, information regarding the use of smokeless tobacco products, such as chewing tobacco or snuff, is also being recorded **(objectives 2 and 4)**.

***Alcohol Consumption:*** Data on alcohol consumption is being collected to assess the drinking habits of the participants **(objectives 2 and 4).**

### Healthcare Access

The following data will be collected from preadolescents and parents.

***Healthcare Needs Assessment:*** Includes an evaluation of the participant’s health status by conducting a physical examination. This involves assessing various aspects of their physical well-being by conducting a clinical examination, assessing vital signs and medical history **(objectives 1, 2 and 3)**.

***Healthcare Utilization***: Information on healthcare utilization is also being collected as part of the study **(objective 4)**. This includes tracking referrals made to healthcare professionals or specialists based on the findings from the physical examination and health history. The study staff follows up with participants to inquire about their utilization of these referrals, including whether they sought the recommended healthcare services, took prescribed medications, and attended follow-up appointments. Additionally, participants are asked to provide information about any relevant biochemical reports they may have received as part of their healthcare utilization. The objective of collecting these data is to assess the participants’ healthcare-seeking behaviours, adherence to medical recommendations, and the effectiveness of the referral system in meeting their healthcare needs.

### Socio-ecological Factors

These data are being collected from the parents.

***Socio-demographic Characteristics:*** The education level and occupation details of the adults and the education level of the primary participant are collected using a questionnaire.

***Socioeconomic Status (objective 3):*** Socioeconomic status (SES) is being assessed using the BG Prasad Scale (45), a widely used classification system for rural populations in India. For the purpose of calculating SES the data being collected comprises the number of members in the household and the total monthly family income. To determine the SES of the enrolled families, we compute the per capita monthly income by dividing the total monthly family income by the total number of members residing in that household. Based on the per capita income of the family, the BG Prasad scale classifies the SES of rural families into the following categories: Upper Class, Upper Middle Class, Middle Class, Lower Middle Class, and Lower Class. To update the BG Prasad Scale for the year 2022, we incorporated the latest Consumer Price Index (CPI) for Industrial Workers (IW) obtained from the Labour Bureau’s website. The CPI for January 2022 was recorded as 125.1. It is important to note that the pilot study was conducted in June 2022. Thus, using the CPI from January 2022 ensured that the socioeconomic classification reflected the most recent economic conditions at the time of enrollment (45).

***Climate Change:*** The impacts of climate change on health behaviours and risk factors are being evaluated using both qualitative and quantitative approaches **(objectives 2 and 3)**. These methods include connecting data on seasonal weather events and variations to dietary habits (i.e., impact of extreme heat or floods on crop yields or types of seasonal food consumed) and health outcomes (i.e., changes in physical activity levels or stress).

***Digital Connectivity and Access:*** Data collected includes participants’ accessibility to and availability of mobile devices and active internet plans which may impact their health-related knowledge, behaviours, and management **(objectives 3 and 4)**.

### Incentives for Study Participation

To encourage participation and as a gesture of appreciation for participants’ time and contributions, appropriate incentives are given to the families. These include monetary compensation for hourly wage lost by participating in the study and non-monetary compensation including healthy snacks at the study site. At enrollment, each family that completes data collection is given a ‘Hygiene Kit’ comprising toothbrushes, toothpaste, bathing soap, detergent soap bar, and hair oil. Incentives are planned to correspond with seasons or events. For example, caps will be given during the summer season, rain gear during the monsoon, and books for the ‘Back to School’ season.

### Data Management, Security, and Analysis Plan

Following data collection, the research assistants check each questionnaire response to reduce the number of missing data entries. The data are entered by trained data entry operators and at regular intervals, the research assistants check the entered data so as to correct data entry errors. Participants (parents and children) and households will be assigned unique study numbers at the time of enrolment. The data will be anonymised. Hence, personal details will only be known to the field team. All members of the field team have signed a Non-Disclosure Agreement and will be trained regularly on the importance of maintaining the confidentiality of the participants. Access to the data will be controlled by the principal investigator with the help of the data manager. Completed questionnaires will be stored in cupboards under lock and a soft copy of the data will be stored on a password-protected computer and a secure cloud server. Both hard and soft copies of data will be accessible only to authorized personnel. While the long-term data analysis plan includes longitudinal modeling, time-series analyses, structural equation modeling, multilevel modeling, and sex and gender-based analyses, this methodological study presents findings from preliminary analyses.

## PRELIMINARY RESULTS

Screening of children started in April 2022 in 20 schools in the 12 selected villages. A total of 2099 children aged 6 to 10 years were screened, of whom 1471 have been found to be eligible for inclusion in the study. Of the 628 children found ineligible for inclusion in the study, a majority (501) would not meet the age inclusion criteria of 8 to 10 years during the study enrolment period, 104 children belonged to migrant families, 22 did not have both biological parents present, and one child had a chronic metabolic condition.

Enrollment started in September 2022. The current rate of enrolment is 12-15 preadolescent boys and girls per week, which is equivalent to 10 families per week. Our team aims to complete the enrollment of 1300 preadolescents (not including parents) by September 2024. With the inclusion criteria necessitating both biological parents of children being included in the study, we expect the total sample size (N), which includes children and their parents to be much larger than 1300. As of 31^st^ May 2023, 378 families have been enrolled, including 432 preadolescents aged 8 to 10 years and 756 parents. The age and gender-wise distribution of enrolled preadolescents are presented in **Table 1**.

**Table 1:**
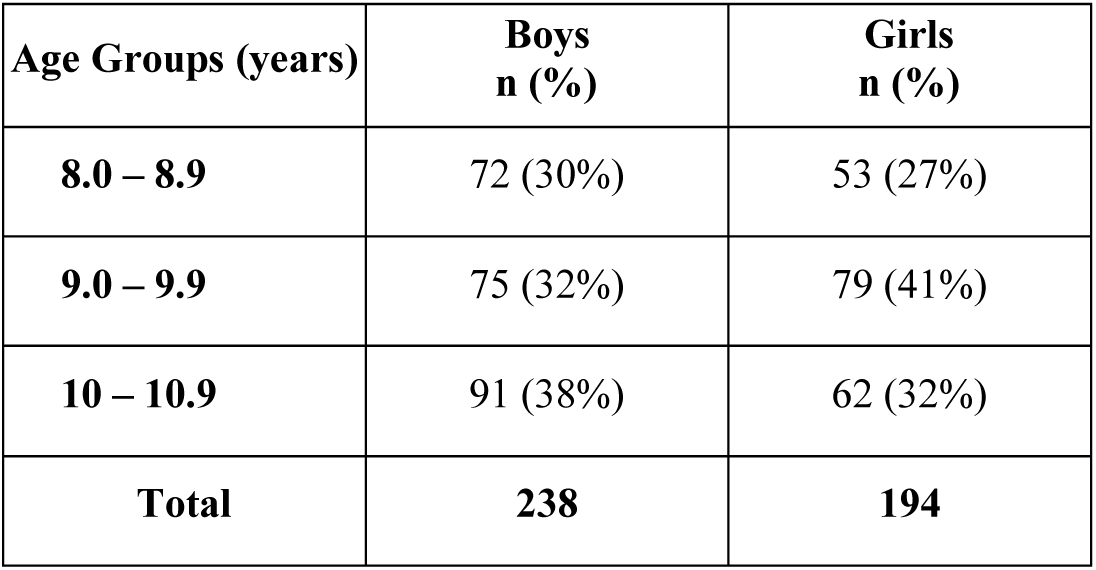
Distribution of Children According to Gender and Age.

| Age Groups (years) | Boys<br>n (%) | Girls<br>n (%) |
| --- | --- | --- |
| 8.0 – 8.9 | 72 (30%) | 53 (27%) |
| 9.0 – 9.9 | 75 (32%) | 79 (41%) |
| 10 – 10.9 | 91 (38%) | 62 (32%) |
| **Total** | **238** | **194** |

The enrolled families were classified into different SES groups using the BG Prasad Scale, considering the latest Consumer Price Index for Industrial Workers obtained from the Labour Bureau’s website (45). Among the enrolled families, the highest SES group (Class I) accounts for 15% of the population. The upper middle class (Class II) represents the largest share at 28%, followed closely by the middle class (Class III) at 32%, indicating a significant portion of the population belonging to these categories. The lower middle class (Class IV) comprises 19% of the population, indicating a relatively smaller proportion compared to the middle and upper middle classes. The lowest SES group (Class V) is the smallest group, representing 7% of the population.

The anthropometric characteristics of the primary participants have been presented in **Table 2**. Approximately 4% of participants were stunted (HAZ < –2.0) and 9% were wasted (WAZ < – 2.0). Based on the Indian Academy of Paediatrics (IAP) classification of BMI, around 12% of children were underweight and 10% were overweight/obese **(Figure 5).**

**Figure 5.**
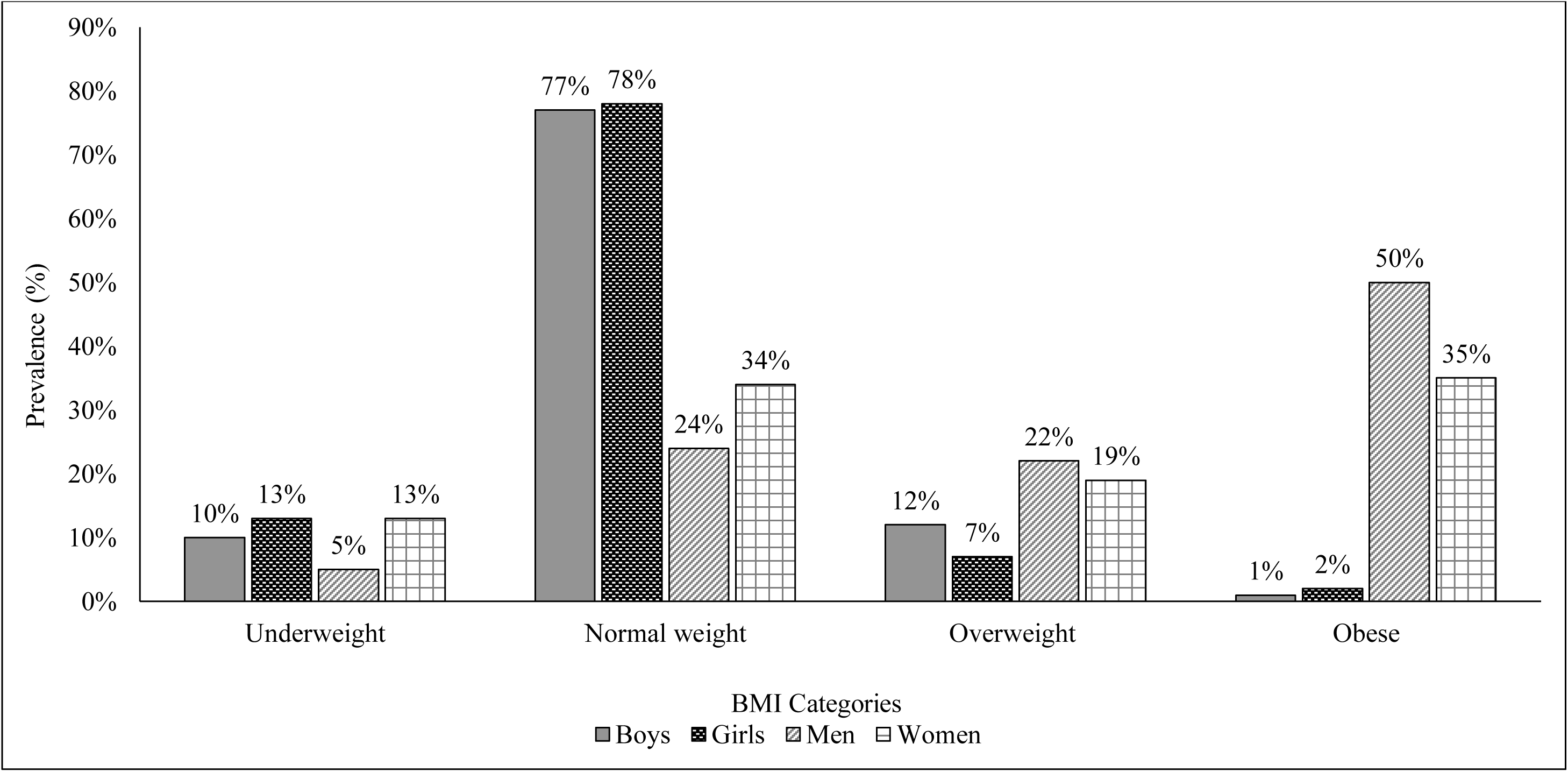
– Nutritional Status of Participants according to BMI categories.

**Table 2:**
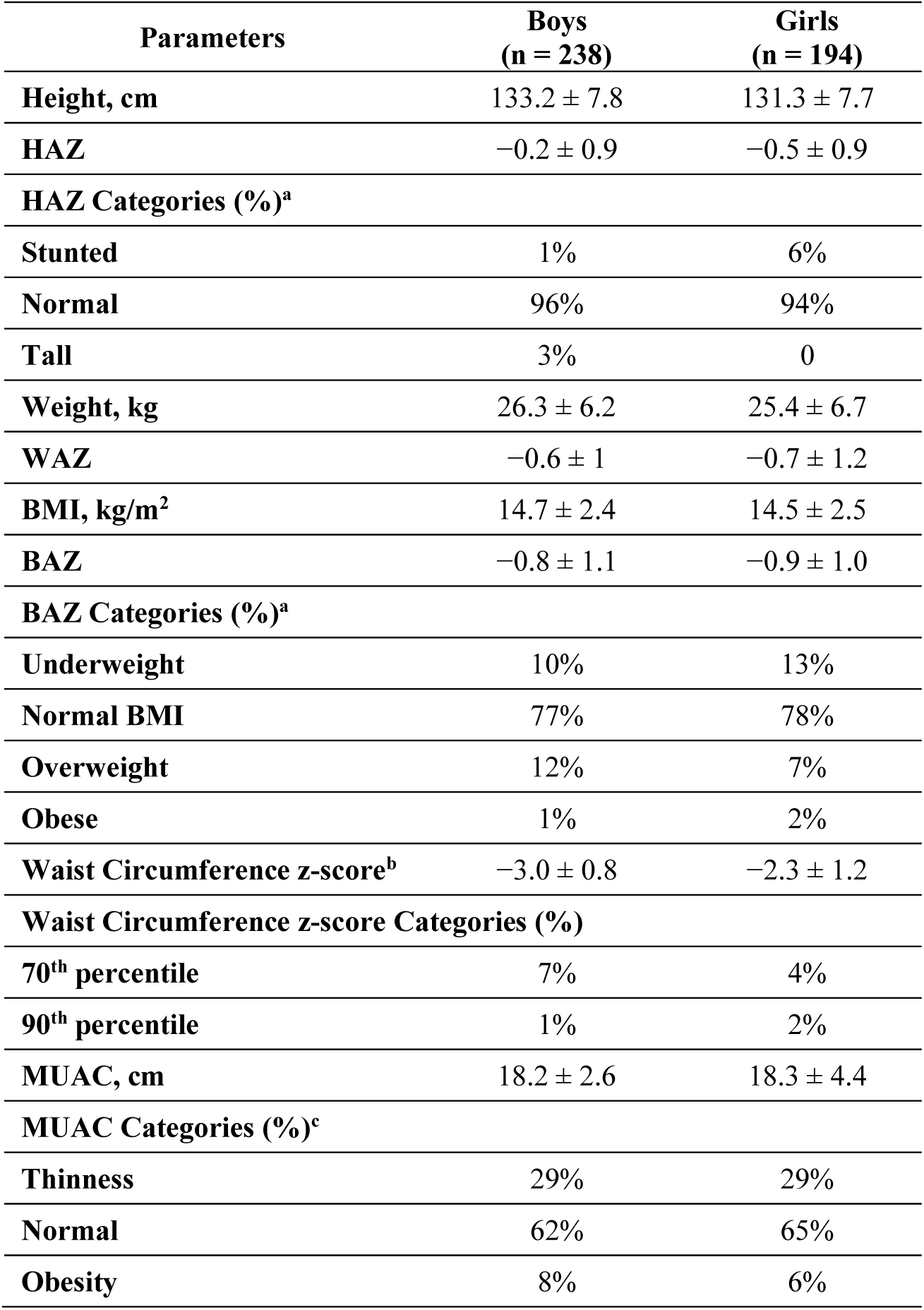
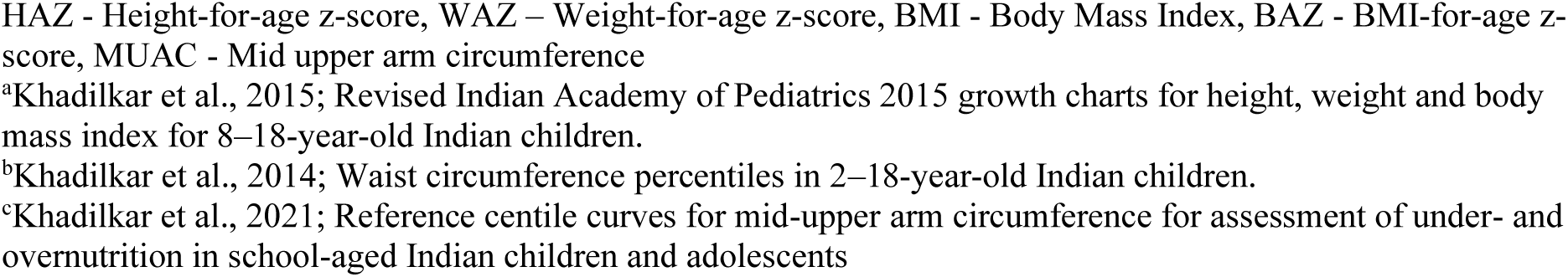
Anthropometric Characteristics and Nutritional Status of the Preadolescents.

| Parameters | Boys<br>(n = 238) | Girls<br>(n = 194) |
| --- | --- | --- |
| Height, cm | 133.2 ± 7.8 | 131.3 ± 7.7 |
| HAZ | −0.2 ± 0.9 | −0.5 ± 0.9 |
| **HAZ Categories (%)<sup>a</sup>** |  |  |
| Stunted | 1% | 6% |
| Normal | 96% | 94% |
| Tall | 3% | 0 |
| Weight, kg | 26.3 ± 6.2 | 25.4 ± 6.7 |
| WAZ | −0.6 ± 1 | −0.7 ± 1.2 |
| BMI, kg/m<sup>2</sup> | 14.7 ± 2.4 | 14.5 ± 2.5 |
| BAZ | −0.8 ± 1.1 | −0.9 ± 1.0 |
| **BAZ Categories (%)<sup>a</sup>** |  |  |
| Underweight | 10% | 13% |
| Normal BMI | 77% | 78% |
| Overweight | 12% | 7% |
| Obese | 1% | 2% |
| Waist Circumference z-score<sup>b</sup> | −3.0 ± 0.8 | −2.3 ± 1.2 |
| **Waist Circumference z-score Categories (%)** |  |  |
| 70<sup>th</sup> percentile | 7% | 4% |
| 90<sup>th</sup> percentile | 1% | 2% |
| MUAC, cm | 18.2 ± 2.6 | 18.3 ± 4.4 |
| **MUAC Categories (%)<sup>c</sup>** |  |  |
| Thinness | 29% | 29% |
| Normal | 62% | 65% |
| Obesity | 8% | 6% |

HAZ - Height-for-age z-score, WAZ – Weight-for-age z-score, BMI - Body Mass Index, BAZ - BMI-for-age z-score, MUAC - Mid upper arm circumference
<sup>a</sup>Khadilkar et al., 2015; Revised Indian Academy of Pediatrics 2015 growth charts for height, weight and body mass index for 8–18-year-old Indian children.
<sup>b</sup>Khadilkar et al., 2014; Waist circumference percentiles in 2–18-year-old Indian children.
<sup>c</sup>Khadilkar et al., 2021; Reference centile curves for mid-upper arm circumference for assessment of under- and overnutrition in school-aged Indian children and adolescents

The age, anthropometric, and clinical characteristics of the parents have been presented in **Table 3**. One in two fathers and one in three mothers were classified as obese based on the World Health Organization Asian-BMI classification **(Figure 5).** Moreover, 22% of fathers and 19% of mothers were classified as being overweight. Contrary to our expectation, only 5% of fathers and 13% of mothers were classified as being underweight. Nearly half of all parents had abdominal obesity with a waist circumference greater than 90 cm in males and 80 cm in females.

**Table 3:**
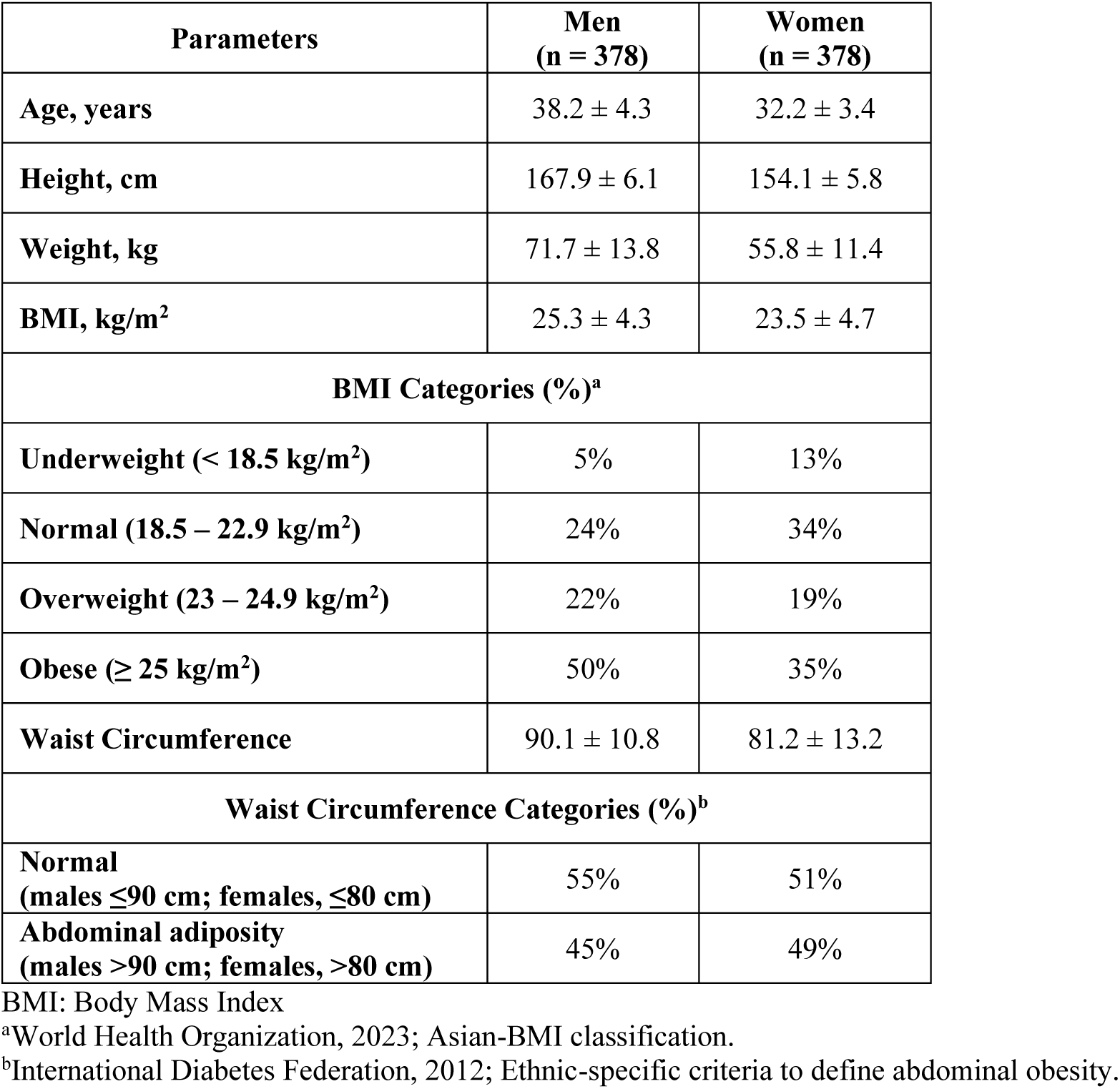
Anthropometric Characteristics and Nutritional Status of the Parents.

The clinical and biochemical parameters of the parents have been presented in **Table 4**. Hypertension was significantly greater among the fathers (26%) as compared to the mothers (7%). One in every four adults had a fasting blood sugar level in the pre-diabetes range of 100 – 125 mg/dL and 5% of adults had type 2 diabetes.

**Table 4:**
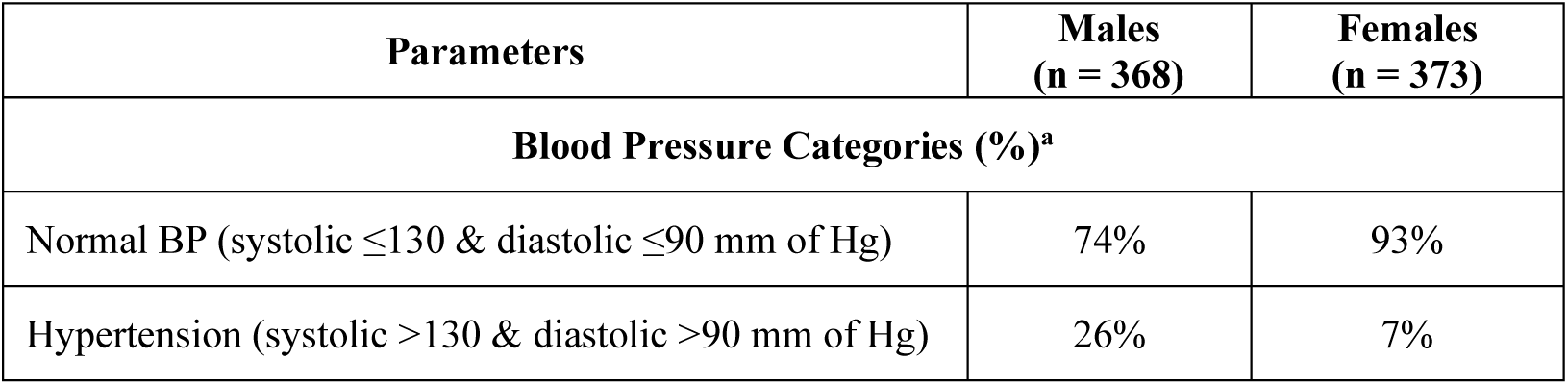
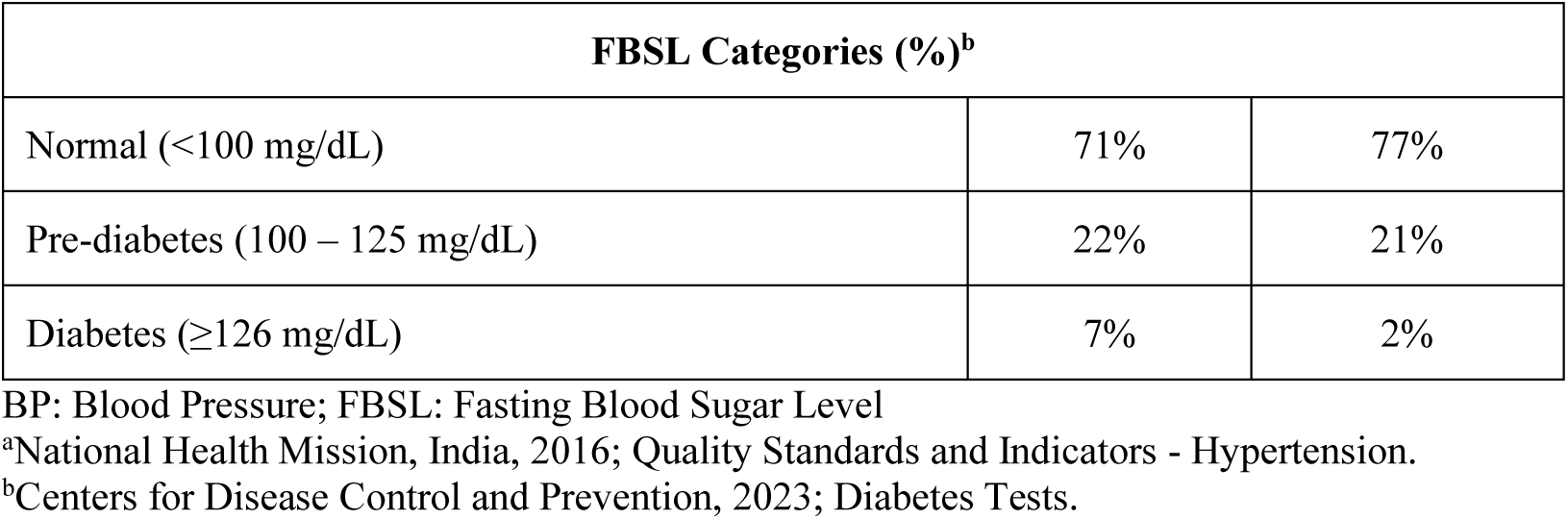
Clinical and Biochemical Parameters of the Parents of the Primary Participants.

The daily physical activity patterns of the participants are described in **Table 5**. Sleep time was the highest for all participants, with a median of 8 hours for adult males, 8 hours for adult females, and 9 hours for children. Following sleep, the participants spent their time differently in terms of physical activity. Adult males and children spent the majority of their time engaged in light physical activity, with median values of 7 hours and 9 hours, respectively. On the other hand, adult females predominantly participated in vigorous physical activity, with a median of 5 hours. Moderate physical activity was also observed, with adult males spending a median of 1-hour, adult females spending a median of 3 hours, and children spending a median of 2 hours. Additionally, the participants’ screen time was recorded, and the data indicated that they spent an average of 2 hours per day on screens **(Figure 6).**

**Figure 6.**
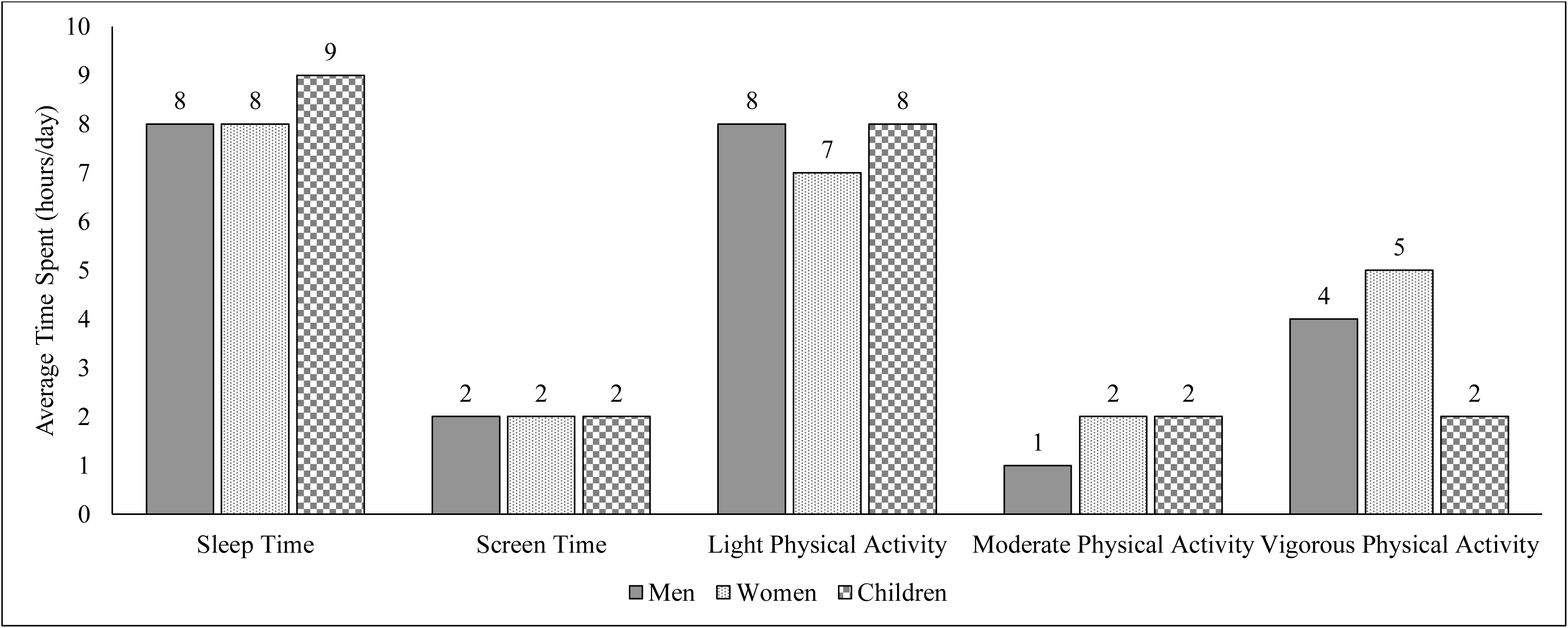
– Physical activity levels among study participants.

**Table 5:** Physical activity patterns of participants (average hours/day)

|  | **Males** |  |  | **Females** |  |  | **Children** |  |  |
| --- | --- | --- | --- | --- | --- | --- | --- | --- | --- |
|  | **Median** | **Percentiles** |  | **Median** | **Percentiles** |  | **Median** | **Percentiles** |  |
|  | **50<sup>th</sup>** | **25<sup>th</sup>** | **75<sup>th</sup>** | **50<sup>th</sup>** | **25<sup>th</sup>** | **75<sup>th</sup>** | **50<sup>th</sup>** | **25<sup>th</sup>** | **75<sup>th</sup>** |
| **Sleep Time** | 8 | 8 | 9 | 8 | 8 | 9 | 9 | 8 | 10 |
| **Screen Time** | 2 | 1 | 3 | 2 | 1 | 2 | 2 | 1 | 3 |
| **Light PA** | 8 | 5 | 10 | 7 | 5 | 9 | 8 | 7 | 10 |
| **Moderate PA** | 1 | 0 | 3 | 2 | 2 | 4 | 2 | 1 | 3 |
| **Vigorous PA** | 4 | 2 | 7 | 5 | 2 | 7 | 2 | 1 | 3 |

## DISCUSSION

The YUVAAN Cohort takes an innovative approach to tracking and addressing multigenerational health behaviours and outcomes among preadolescents and their parents in India. Taking a life course perspective, this is the only study to date which enables understanding of the complex socio-ecological determinants of health that influence growth trajectories and intergenerational NCD risk among rural communities – many of which experience greater vulnerability to low socio-economic status, varied healthcare access, and increasingly, the brunt of adverse climate change impacts (46). As India contributes a significant proportion of the global workforce (47), the burden of NCDs will impact the health, social, and economic well-being of society at home and abroad, thus requiring dedicated focus, particularly among the most vulnerable groups.

As of May 31, 2023, 378 families have been successfully enrolled into the YUVAAN Cohort, comprising 432 preadolescents and 756 parents. Going forward, data will be collected from future spouses and children of the enrolled preadolescents, thereby expanding the data collection to three generations. This holistic approach allows for a comprehensive understanding of intergenerational health risks, as well as the potential to break the cycle of NCDs within families and communities.

Based on preliminary results, the YUVAAN Cohort study has identified obesity to be a concern among households in rural areas. This finding is in line with previous studies examining the shifting burden of obesity incidence, demonstrating the increased prevalence of obesity in rural areas and lower to middle socioeconomic groups (48,49). Paradoxically, these findings also highlight the double burden of malnutrition, with undernutrition and overweight/obesity coexisting among children and parents. This observation is consistent with earlier studies conducted in India and other low and middle-income countries like Brazil and Mexico, that have documented the coexistence of different forms of malnutrition within populations (50–52).

Comparing our initial results to those of previous studies, it is evident that there have been changes in the prevalence and patterns of NCDs over time (53). Preliminary findings from our study identifies high rates of diabetes and hypertension among men and women in rural areas, surpassing the figures reported in previous research conducted in similar regions. This discrepancy suggests that there may have been significant shifts in the burden of these diseases within the Indian population, where traditionally the burden of these NCDs has been seen predominantly in urban cohorts (53). These findings align with previous studies that have highlighted the impact of urbanization-induced characteristics in the food and physical environments among rural areas, which promote obesity, diabetes, and other cardio-metabolic diseases (54). The transition from traditional lifestyles to more sedentary and unhealthy behaviours associated with urbanization may also explain the observed patterns (55). Based on the dietary data collected, our initial observations indicate that the consumption of bakery products like breads, khari (puff pastries), and biscuits is quite frequent, sometimes replacing meals like breakfast for children and adults. We also note frequent consumption of ultra-processed foods like chips, and noodles. Although these findings will be explored going forward and addressed as necessary, it is important to note that the increasing threat of obesity among the rural poor is particularly concerning, as it increases the burden and severity of NCDs (56,57).

In terms of the overall approach and focus of implementation, where previous studies have primarily focused on documenting the prevalence of NCDs (53,55,57), the YUVAAN Cohort study takes a more holistic approach by studying growth trajectories with the aim of breaking patterns of intergenerational transmission of health risk. This approach involves a shift from participant observation to interventions where possible – an ethical approach in global health to provide communities with necessary healthcare and support (58). Moreover, this comprehensive approach will allow for a deeper understanding of the complex dynamics involved in the development and prevention of NCDs across generations, particularly among vulnerable rural populations.

It is also important to note that this study is taking place during the age of climate emergency (59,60). In addition to the cumulative risk factors that rural families face for NCDs, climate change-related events – including increasingly frequent and severe heat waves, droughts, and extreme storms (59) – further burden NCD prevention and management. Thus, it is critical to not only take a life course perspective in this work, but also a systems thinking lens which captures the complexity of socio-ecological determinants of health (60). As a result, the YUVAAN Cohort study will explore the impacts of climate change on NCD risk, and work to integrate advanced digital epidemiological approaches to improve data collection processes for both objective climate-related data, as well as health behavioural data (61–63).

Systems thinking takes a non-linear perspective to disease pathways, and provides a framework for understanding reciprocal relationships and interactions between variables within systems (e.g., food access – food production within food systems), and across systems (e.g., between food access and nutrition across health and food systems) (60). This approach is inherently symbiotic not only with the complexity of NCD risks, but also critical in understanding and addressing interdependent risks of NCDs in the age of climate emergency. Global population health crises in the 21^st^ century are extremely complex, with links to economic disasters (64–66), warfare (64), and climate change (64,66). Nevertheless, the digital age offers new opportunities and challenges to tackle these global crises (67).

For instance, digital tools and technologies are increasingly being used to not only address urgent humanitarian crises (68) but also facilitate citizen participation, health system, and population health interventions, and knowledge transfer (68–71). Given that addressing intergenerational transmission of health inequities is one of the primary goals of the YUVAAN study, there is a role for digital tools and technologies in contributing to the reduction of health disparities, where citizen-owned digital tools can be used to amplify their voices (71,72).

As identified during enrollment, the vast majority of YUVAAN study participants own smartphones, which corroborates existing evidence on the penetration of these ubiquitous tools in India (73). Thus, we intend to ethically leverage citizen-owned smartphones to not only improve our longitudinal data collection and compliance, but also the implementation and evaluation of potential interventions. This approach can aid in minimizing loss to follow-up in longitudinal cohorts (74), improve adherence of interventions (75), and perhaps more importantly, enable real-time support to the most vulnerable populations (70,77).

There are several key strengths of the YUVAAN cohort study. First, this study is being conducted through an Indo-Canadian partnership, which involves combining interdisciplinary expertise in population health, health systems, healthcare, digital epidemiology, and community-based participatory research. This partnership focuses on equity in collaboration and capacity building over time, which is critical for research being conducted with vulnerable populations in the global south (77). Moreover, the dynamic expertise of the team and diversity of data collected will enable targeted interventions in the future (i.e., randomized clinical trials, natural experiments, and smartphone-based digital health interventions) tailored to the specific health needs and concerns of rural populations.

However, given the longitudinal nature of this cohort study, we anticipate numerous challenges including compliance and retention across generations, and continuous communication with multiple villages as enrollment increases. These challenges highlight the need for sustained community relationships and a shift to digital data collection to enable remote communication and accessibility for long-term study participation. There are also potential limitations with several of the measures used. For example, while BMI will be computed it does not capture abdominal adiposity which is considered to be riskier for specific NCDs (78). However, the use of several clinical measures and objective data collection tools for growth, body composition using the DXA, bone density, muscle strength and function, and biochemical parameters will minimize reliance on single variables to determine health behaviours and associated risks. Also, the use of the 24-hour dietary recall method and physical activity questionnaire may lead to inaccuracies in the information collected, so we are collecting dietary data over three non-consecutive days using the multiple-pass method and food models to aid in accurate recall and minimize bias and using a validated questionnaire for physical activity.

In conclusion, the YUVAAN Cohort study will contribute significantly to the progress in investigating and understanding the intergenerational transmission patterns of NCDs in rural India. The innovative study design, multi-generational perspective, and socio-ecological approach will enhance our understanding of NCD prevalence, prevention, and the impact of various factors on health outcomes. The findings of this study have implications for public health practice, including expanding our understanding of complex health systems and associated interventions and policies which will be necessary to address the specific health needs and concerns of rural populations. By breaking the cycle of NCDs within families and communities, the study aims to improve the overall health and well-being of rural communities in India and contribute to the global efforts in combating NCDs.

## Data Availability

All data produced in the present study are available upon reasonable request to the authors

## Notes

### Competing Interest Statement

The authors have declared no competing interest.

### Clinical Protocols

https://classic.clinicaltrials.gov/ct2/show/NCT05603793

### Funding Statement

This study did not receive any funding

