## Supplementary Data for "The YUVAAN cohort: an innovative multi-generational platform for health systems and population health interventions to minimize intergenerational transmission of non-communicable diseases in India"

**Anthropometric Measurements:** In adults and children, standard protocols are being used to measure the standing height and sitting height by a portable stadiometer (seca 213 Portable Stadiometer; seca, Hamburg, Germany) to the nearest 0.1 cm and body weight using the Tanita Body Composition Analyzer (Model MC-780 MA; Tanita Corporation of America, Arlington Heights, Illinois). Body mass index is computed as weight in kilograms divided by height in meters squared and is expressed as kg/m2. Anthropometric Z-scores for children are being computed using ethnic-specific growth references (41). Additionally, we are measuring the waist circumference (WC) in both adults and children in accordance with the method used in the National Health and Nutrition Examination Survey (NHANES) III. WC is being measured with the participant standing using a stretch-resistant tape applied horizontally just above the upper lateral border of the right ileum. Each measurement is made at the end of a normal expiration and is recorded to the nearest 0.1 cm. Mid-upper arm circumference (MUAC) is being measured only in the pre-adolescent at the mid-point between the acromion and the olecranon processes using a stretch-resistant Seca tape.

Bone age - Bone age assessment is performed for skeletal maturity. For the purpose of bone age assessment x-rays of the left hand and wrist were performed by exposing them in posteroanterior (PA) position with the elbow and wrist on the same axis, thumb at an angle of 30°, and fingers not too close or widely spaced. The X-ray was centred overhead of the third metacarpal bone from a distance of 76 cm. The radiographic examinations were performed using a portable X-ray generator machine (Philips, Eindhoven, Netherlands) operating at 50 mA and the digital X-rays were generated by the standard computed radiography system (Fuji, Tokyo, Japan). Each x-ray was rated by the same observer using the Tanner Whitehouse-3 method.

**Biochemical Assessments:**

Biochemical assessments for children include estimation of haemoglobin concentrations, blood sugar levels, and indicators of bone metabolism and growth modulators such as serum calcium, phosphate, alkaline phosphatase, parathyroid hormone (PTH), vitamin D, and luteinizing hormone (LH). Biochemical assessments for parents include the estimation of haemoglobin concentrations, blood sugar levels, and lipid profiles. Blood for biochemical estimations is collected in a fasting state by trained phlebotomists at the residence of the participants. Venous blood is being collected in BD Vacutainer blood collection tubes -  lavender (EDTA) tubes for haemoglobin estimation and gold (gel-containing) tubes for all the other tests. Blood samples collected in Lavender tubes are stored at room temperature and processed within 24 hours on the Horiba Yumizen H500 for haemoglobin estimation by spectrophotometry. Blood samples collected in the gold tubes are centrifuged at 3000 rpm for 15 minutes at room temperature within 2 hours of collection. Serum calcium, phosphorus, creatinine, alkaline phosphatase and lipid profile (for parents) are estimated on the Selectra Pro S Automatic Biochemistry Analyzer using the QLine BioTech (India) Rapid Test Kit within 24 hours of sample collection. Serum Luteinizing hormone is estimated by the Chemiluminescent Microparticle Immunoassay (CMIA) method using the ARCHITECT LH Reagent Kit (6C25)). Serum aliquots are being stored at -20°C for future estimations of serum parathyroid hormone and vitamin D.  Serum 25(OH)D concentrations will be assessed by enzyme-linked immunosorbent assay (ELISA) using standard kits (DLD Diagnostika GmbH [Hamburg, Germany]; intra-assay coefficient of variation [CV]: 5%; interassay CV: 7.8%). Serum intact PTH concentration too will be assessed by the ELISA technique using standard kits (Biomerica Inc [Irvine, California]; sensitivity: 0.17 pmol/L; interassay CV: <4%; intra-assay CV: 3%-6%).

**Body Composition Assessment:**

The MC-780 model of the Tanita Body Composition Analyzer was used to measure body weight and assess body composition using bioelectric impedance analysis (BIA). Participants are instructed to step onto the platform with their feet touching all four metal plates following which they are instructed to hold the hand grips. The instrument measures body composition as fat percent and mass, fat-free mass, muscle mass, protein, body water and bone mineral mass.

**Bone Parameters Assessment:**

- Dual X-ray Absorptiometry (DXA) - The GE Lunar iDXA (Wisconsin, MD) (Software encore version 16) was used to measure bone density in the anteroposterior (AP) spine and assess total body composition. Bone Mineral content (BMC), Bone Area (BA) and areal Bone Mineral Density (aBMD) are being measured in the L1-L4 region. The participant is instructed to lie on their back in the centre of the iDXA scanner table with their arms along their side. The legs of the participant are elevated using a foam block leg positioner to achieve an angle of 60° to 90° between the patient’s thighs and the scanner table so as to separate the vertebrae and flatten the lower spine. The AP spine is scanned from the top of the L5 vertebra to half of the T12 vertebra. The CV for L1 - L4 aBMD and L1 - L4 BMC for standard analysis are 1% and 2.8%, respectively. The DXA software typically performs the analysis automatically after the completion of the scan by using the machine-generated Z-scores at L1 - L4 aBMD. Calibration of the machine is done daily. The same technician performs all the scans using the same scan mode which is automatically selected.

Body composition was measured by assessing total body less head. The iDXA measures total percent body fat, total Body Fat Mass (g), Fat free Mass (FFM) (g) and Bone mineral content (BMC) (g).

- Peripheral Quantitative Computerised Tomography (pQCT)  - The pQCT measurements are being  performed at the radius of the non-dominant hand using the Stratec XCT 2000 equipment (Stratec Inc., Pforzheim, Germany). These are analysed by the integrated software for Stratec 2000, version 6.2. First, a non-stretchable tape is used to measure the distance between the ulnar styloid process and the olecranon process on the nondominant arm of the participant. pQCT images are then taken at lengths of 4% and 66% of the radius by employing the scout view to draw a reference line through the middle of the ulnar border of the articular cartilage (F Rauch 2005, 2008). Trabecular volumetric BMD (vBMD) and bone mass are measured at a 4% radius at a threshold of 180 mg/cm^3^ using 0.59 mm voxel size, slice thickness of 2.5 mm, contour mode 2, and peel mode 2. At 66% of the length of the radius, using a threshold of 711 mg/cm^3^ and contour mode 3 the other pQCT parameters including cortical vBMD, cortical thickness, periosteal circumference, endosteal circumference, and polar strength strain index (SSI) are measured. The CV for measurement of total, trabecular and cortical density are 0.7, 3.0, and 0.8% respectively. The CVs for cortical thickness, periosteal circumference and endosteal circumference measurements are 2.6%, 0.8% and 1.9%, respectively. Movement artefacts are  assessed by visual inspection by the same operator and if the quality of the scan was poor or degraded the scan was repeated. For parameters like trabecular density, total density, cortical density, SSI and muscle area for height Z-scores for age generated by the pQCT machine will be used for analysis.
- Jumping Mechanography (JM) - Dynamic muscle function, for children and adults, is being assessed using the Leonardo Mechanograph Ground Reaction Force Plate (Novotec Medical, Pforzheim, Germany). The software provided by the manufacturer (Leonardo Mechanography GRFP version 4.4, Novotec, Pforzheim, Germany) is being used for the calculation of the muscle function outcomes. All the participants are being instructed to perform two types of jumps: single 2-legged jump (s2LJ) which detects the maximum relative power and multiple 1-legged hopping (m1LH) which detects the maximum relative force (Kasture et al., 2022). The technician has been trained to demonstrate the jumps to the participants and she also gives instructions as the participant is performing the jumps. The participants are instructed to repeat each type of jump until 3 acceptable jumps are obtained. The jump with the greatest peak power/force will be used for analysis. In order to perform the s2LJ participants are instructed to squat briefly before jumping so that the s2LJ can be performed as a counter-movement jump with freely moving arms. The maximum power relative to body mass (Pmax/mass, Watt/kg) is the main outcome of interest for the s2LJ. Apart from Pmax/mass, this test also generates the Esslinger Fitness Index (EFI) outcome, which is the maximum power relative to body mass normalised to age and gender, and the standard deviation score of EFI (EFI-SDS). To perform m1LH participants are instructed to hop repeatedly and as fast as possible on the forefoot of their dominant leg (approximately fifteen jumps). The manufacturer’s software automatically excludes any repetition in which there is heel contact from the analysis. The main outcome obtained from the m1LH is the maximum relative force i.e. Fmax normalised to body weight (Fmax/BW). The inter-day test-retest measurements of the main outcome parameters i.e. peak force relative to body weight or peak power relative to body weight have shown low variability (3.4% to 7.5%) in healthy children and adults (Veilleux & Rauch, 2010).
- Hand Grip Dynamometer - The grip strength of the non-dominant hand was measured for all participants using the Jamar® Plus+ Digital Hand Dynamometer (Patterson Medical, Warrenville, IL, USA) as per standard procedure protocol (Ewing Fess & A. Moran, 1992). Each participant is instructed to sit in an upright position on a chair (without an armrest), with the shoulders adducted and the feet flat on the ground. Prior to performing the measurement, the participants are given a demonstration on how to perform the test by a trained technician. The dynamometer is adjusted as per the size of the participants’ hand. The participant is instructed to hold the dynamometer in the non-dominant hand and to keep the arm at an angle of 90º at the elbow, with the forearm and wrist in a neutral position and the thumb facing upwards. The elbow remains unsupported during the test. The technician verbally encourages each participant to squeeze the dynamometer as hard as possible so as to generate the maximum possible effort. An average of three readings is taken as the grip strength (kg) of the participant (Coldham et al., 2006). Additionally, we are calculating ‘relative grip strength’ to adjust for the effect of body weight (Grip strength/body weight; GS/kg (no unit)).
