## Supplementary Table 6 for "The YUVAAN cohort: an innovative multi-generational platform for health systems and population health interventions to minimize intergenerational transmission of non-communicable diseases in India"

**Table 6 -** Detailed timeline of data collection and assessments (For young adolescents aged 8-10 years)

| **Data** | | **Timeline of measurement** | | | **Measuring Tool** |
| --- | --- | --- | --- | --- | --- |
|  |  | **Baseline** | **Every**  **6-months** | **Every**  **1-year** |  |
| **1** | **Personal Details & Socio-demographic (Geomapped)** | x |  | x | Questionnaire & BG Prasad scale |
| **2** | **Medical History** | x |  |  | Clinical Questionnaire |
| **3** | **Birth Weight** | x |  |  |  |
| **4** | **Clinical Assessment** | x |  | x | Clinical Questionnaire |
| **5** | **Anthropometry** |  |  |  |  |
|  | Height | x | x | x | Harpenden stadiometer |
|  | Weight | x | x | x | TANITA MC780 MA |
|  | Sitting Height | x | x | x | Harpenden Stadiometer and a flat-surfaced seat, 50 cms high standard chair |
|  | Mid-Upper Arm Circumference  (MUAC) | x | x | x | Non elastic measurement tape |
|  | Waist Circumference | x | x | x | Non elastic measurement tape |
| **6** | **Biochemical Parameters** |  |  |  |  |
|  | Haemoglobin | x |  | x | Horiba Yumizen H500 |
|  | Fasting Blood Sugar (glucose) Level | x |  | x | Glucometer |
|  | Alkaline Phosphatase | x |  | x | Selectra Pro S |
|  | Sr. Calcium | x |  | x | Selectra Pro S |
|  | Sr. Phosphorous | x |  | x | Selectra Pro S |
|  | S. Vitamin D (25 OH-D) | x |  | x | ELIZA |
|  | S. Creatinine | x |  | x | Selectra Pro S |
|  | Luteinizing Hormone (LH) | x |  | x | Chemiluminescent Microparticle Immunoassay (CMIA) |
|  | Parathyroid Hormone (PTH) | x |  | x | ELIZA |
| **7** | **Substance Abuse** | x |  | x | Questionnaire |
| **8** | **Physical Activity** | x |  | x | Questionnaire |
| **9** | **Sunlight Exposure** | x |  | x | Questionnaire |
| **10** | **Dietary Recall** | x |  | x | Questionnaire |
| **11** | **Bone Health & Muscle Function** |  |  |  |  |
|  | Bone Age | x |  | x | Skeletal x-ray |
|  | Bone Mineral Content | x |  | x | Lunar iDXA |
|  | Bone Mineral Density | x |  | x | XCT 2000 (pQCT densitometer ) |
|  | Muscle Function | x |  | x | Leonardo Jumping Platform (Jumping Mechanography) |
|  | Grip Strength | x |  | x | JAMAR Plus+ Digital Hand Dynamometer |
