## Supplementary Table 7 for "The YUVAAN cohort: an innovative multi-generational platform for health systems and population health interventions to minimize intergenerational transmission of non-communicable diseases in India"

**Table 7 -** Detailed timeline of data collection and assessments (For adults)

| **Data** | | **Timeline of measurement** | | | **Measuring Tool** |
| --- | --- | --- | --- | --- | --- |
|  |  | **Baseline** | **Every 2-  years** | **Every 5-  years** |  |
| **1** | **Personal Details & Socio-demographic (Geomapped)** | x | x |  | Questionnaire & BG Prasad scale |
| **2** | **Medical History** | x | x |  | Clinical Questionnaire |
| **4** | **Clinical Assessment** | x |  |  | Clinical Questionnaire |
| **5** | **Anthropometry** |  |  |  |  |
|  | Height | x | x |  | Harpenden stadiometer |
|  | Weight | x | x |  | TANITA MC780 MA |
|  | Sitting Height | x | x |  | Harpenden Stadiometer and a flat-surfaced seat, 50 cms high standard chair |
|  | Waist Circumference | x | x |  | Non elastic measurement tape |
| **6** | **Biochemical Parameters** |  |  |  |  |
|  | Haemoglobin | x | x |  | Horiba Yumizen H500 |
|  | Fasting Blood Sugar (glucose) Level | x | x |  | Glucometer |
|  | Lipid Profile | x |  | x | ELIZA |
| **7** | **Substance Abuse** | x | x |  | Questionnaire |
| **8** | **Physical Activity** | x | x |  | Questionnaire |
| **9** | **Sunlight Exposure** | x | x |  | Questionnaire |
| **10** | **Dietary Recall** | x | x |  | Questionnaire |
| **11** | **Bone Health & Muscle  Function** |  |  |  |  |
|  | Bone Mineral Content | x | x |  | Lunar iDXA |
|  | Bone Mineral Density | x | x |  | XCT 2000 (pQCT densitometer ) |
|  | Muscle Function | x | x |  | Leonardo Jumping Platform (Jumping  Mechanography) |
|  | Grip Strength | x | x |  | JAMAR Plus+ Digital Hand Dynamometer |
