## Supplementary Table 8 for "The YUVAAN cohort: an innovative multi-generational platform for health systems and population health interventions to minimize intergenerational transmission of non-communicable diseases in India"

**Table 8 -** Detailed timeline of data collection and assessments (For offspring born to primary participants)

| **Data** | | **Timeline of measurement** | | **Measuring Tool** |
| --- | --- | --- | --- | --- |
|  |  | **Baseline** | **Every 1 year till neonate turns 5 years** |  |
| **1** | **Personal Details & Socio-demographic (Geomapped)** | x | x | Questionnaire & BG Prasad scale |
| **2** | **Clinical Assessment** | x | x | Clinical Questionnaire |
|  | Birth Weight | x |  |  |
| **3** | **Anthropometry** | x | x |  |
|  | Length/ Height | x | x | Infantometer |
|  | Weight | x | x | Digital Weighing Scale |
|  | Head Circumference | x | x | Non elastic measurement tape |
|  | Mid-Upper Arm Circumference (MUAC) | x | x | MUAC Tape |
| **4** | **Infant & Child Feeding Patterns** | x | x | Questionnaire |
| **5** | **Bone & Body Composition** | x | x | Lunar iDXA |
